# A Personalized, Symptom-Based Approach to Boost Research Participation in Hospitalized Older Adults

**DOI:** 10.64898/2026.07.12.26357874

**Authors:** Nicole Ceriani, Shreya Dhar, Catherine Zhao, Ian Sherrington, Eyal Y. Kimchi

## Abstract

**Background:** Delirium is common among hospitalized older adults on many clinical services and associated with poor outcomes. Given delirium’s fluctuations, wearable devices are promising continuous monitors. While recruiting for a wearable electroencephalography (EEG) delirium study, we initially experienced low enrollment rates among older adults and patients on non-neurologic services. Our aim was to understand patient and community perspectives on inpatient, wearable research to adapt recruitment protocols and increase enrollment.

**Methods:** We approached patients admitted to an academic medical center to participate in an observational, wearable EEG delirium study and recorded reasons for enrolling or declining. To gain insight into recruitment protocols, we held a community panel with patients, family members, and caregivers. Recruitment protocols were refined in two phases: 1) personalizing the recruitment approach to emphasize symptoms that were personally relevant to individual patients and 2) sharing educational materials about the study in addition to delirium. We compared enrollment rates before and after these protocol adaptations.

**Results:** Initially, 18.5% of approached patients enrolled (68/367). Despite antecedent concerns that wearable devices would be the primary deterrent to participation, only a small proportion of people who did not participate did so because of wearable EEG (8.8%, 26/299). Community panel members (n=7) suggested that personal relevance and understanding of the clinical conditions being studied, such as delirium, would have a greater impact on decisions to participate than study procedures. Adapting recruitment protocols to highlight personally relevant delirium-related symptoms, such as sleep disturbance, significantly increased enrollment rates (30.1%, 58/188, p<0.001), including for patients over 65 years old (p<0.001) and patients on non-neurologic services (p<0.001). The addition of educational materials focused on clinical delirium did not further impact enrollment (p=0.61).

**Conclusions:** Recruitment of older, hospitalized patients for inpatient research can be challenging, but can be significantly improved by highlighting familiar symptoms of personal relevance.

## Introduction

Hospitalized older adults are at increased risk of developing multiple clinical complications including delirium, an acute and fluctuating disturbance in attention and awareness associated with morbidity and mortality.^1,2^ Delirium occurs due to a multifactorial mix of predisposing and precipitating causes, and can occur on all clinical services. Given the lack of FDA-approved therapies for delirium and the challenges of broadly implementing multifaceted, non-pharmacologic programs, prospective research into delirium is necessary to improve targeted and efficient care. However, older adults, who are at highest risk of developing delirium, may decline clinical research participation at more than double the rate of younger individuals.^3^

Older adults may be less likely to participate in clinical research due to several reasons, including increased appraisal of study components as intrusive,^4^ accrued suspicion of research over their lifetime,^4^ and discouragement of participation by protective relatives.^3^ Older patients do, however, continue to participate in clinical research studies for personal health benefit.^5^ Despite the widespread clinical relevance of delirium to older adults, general knowledge of this canonical geriatric syndrome remains surprisingly limited.^6^ Additionally, the variety of delirium symptoms, including but not limited to fluctuating confusion and sleep disruption, may have different personal significance for different people.^7^ Older adults therefore may not immediately appreciate or identify with the personal relevance of delirium research, especially in an acute and dynamic clinical context.

The dynamic and fluctuating nature of delirium suggests that tools that can monitor physiology continuously, such as EEG, may improve monitoring or understanding of delirium.^8,9^ Continuous clinical EEG, while predictive of delirium,^10,11^ can be cumbersome and restrictive for older patients with acute medical illness. In contrast, wearable EEG can be less restrictive and applied more broadly than clinical EEGs,^12,13^ but older adults who are most at risk of delirium are less likely to use wearable devices.^14,15^ Additionally, familiarization with devices is important for older adults considering the use of one,^16^ and patients with non-neurologic health concerns may not be familiar with EEG devices.

We developed a prospective, observational study spanning multiple medical services and ages to characterize how people’s brains respond during hospitalization, including delirium (Study on Acute Brain Resilience, SABRE). However, enrollment rates were less than anticipated amongst older adults and patients on non-neurologic services. We therefore carried out a broader investigation of attitudes and opinions on delirium and wearables research. Our aim was to understand what factors may influence participation in an acute observational study of a geriatric syndrome such as delirium. Here, we report insights from patients and community panel members that led to changes in our recruitment protocols, whose efficacy we tested. Our findings provide practical recommendations to enhance the participation of older patients in acute clinical research, primarily through personalized recruitment conversations.

## Methods

### Study Design and Participants

We examined enrollment data from SABRE, a prospective, observational study that uses wearable EEG for continuous monitoring and repeated cognitive assessments to assess for delirium during a hospitalization, from enrollment to discharge. The purpose of the study as described on the patient consent form was as follows: “The purpose of this research is to determine how people’s brains respond during hospitalization. Hospitalization can be challenging, but everyone responds differently, for example some people develop difficulty sleeping, others may develop confusion. Understanding how many people’s brains respond during hospitalization may lead to treatments to help prevent hospital-acquired confusion and delirium, maximize brain resilience during hospitalization, and improve sleep and recovery during and after illness.”

Required components of the study included up to twice daily clinical assessments of delirium along with wearable EEG. Optional study components included continuous video recording to capture behavioral features not fully characterized by EEG, and repeated blood sampling to assess potential biomarkers. Participation in any study component was voluntary and optional elements could be declined independently of the core study procedures (EEG and clinical assessments) without affecting enrollment or participation in other study activities. Patients or their legally authorized representatives (LARs) could withdraw from the study at any time without affecting the patient’s clinical care.

Inclusion criteria included adult patients 18 years or older admitted to an academic medical center (Northwestern Memorial Hospital) for at least one night. Exclusion criteria included non-English speakers (due to the language-specific nature of cognitive assessment scales used) and patients with scalp incisions or head wounds that would prevent EEG device application. The study was approved by the Northwestern University Institutional Review Board (protocol STU00218518).

### Recruitment Process

A study team member screened inpatient census lists daily and reviewed patients’ eligibility through the electronic medical record to enroll patients within 48 hours of admission, without interrupting clinical care. Screening and recruitment prioritized patients with evidence of active infection or high clinical suspicion of infection identified in the medical records, which included fever, antibiotic use, or diagnoses suggestive of systemic or focal infection. In particular, patients with infections such as pneumonia or urinary tract infection were prioritized due to their established association with delirium.^17,18^ We intentionally approached a balanced mix of patients who were already experiencing acute altered mental status and those at elevated risk but not yet delirious, allowing observation of delirium onset, fluctuations, and recovery during hospitalization.

Once an eligible patient had been identified, we contacted the nursing staff for permission to approach the patient. Patients were approached in person for study recruitment. A research team member first introduced themselves as research staff and confirmed with the patient that it was an appropriate time for a five-minute discussion regarding the study. Research staff assessed decision-making capacity during this interaction, which included the patient’s ability to understand and engage with study-related information, supplemented by input from the patient’s nurse. If the patient lacked decision-making capacity at the time of consent, a study team member provided a detailed explanation of the study to the patient’s LAR, showing the wearable EEG device during the encounter (see Supplementary Material Figure S1). When LARs were present at the patient’s bedside, consent was obtained in person; if not physically present, LARs were contacted by telephone and provided consent electronically via a REDCap-based e-consent form distributed by email or text message. We tracked decisions to participate or decline (starting September 5, 2023) and primary reasons for those decisions (starting on November 6, 2023, and ending on February 6, 2025).

### Demographic and Clinical Data

Demographic data were extracted from the medical record. We also extracted the presence of comorbidities prior to the date of admission and admission diagnoses based on International Classification of Diseases, 10th Revision (ICD-10) codes. Baseline cognitive impairment was defined as the presence of either Mild Cognitive Impairment or Dementia codes as of the date of admission. We additionally extracted anonymized, demographic and diagnostic data from all patients admitted to the hospital during the study period who met retrospectively verifiable inclusion criteria (18 years or older, at least one night’s stay, English speaking). We excluded patients admitted to Obstetrics and Behavioral Health wards as we did not attempt to recruit such patients in the prospective study.

### Community Panel

To gain additional insight into attitudes about inpatient delirium and wearable research, we conducted a Shared Resource Panel (SHARP), with assistance from Northwestern Medicine’s Center for Community Health,^19^ to facilitate a virtual panel discussion over Zoom with patients, family members, and caregivers. The panel was advertised as addressing “brain health, sleep quality, and alertness during and after hospitalization” and called for individuals who had either been hospitalized or served as a primary caregiver within the past five years. During the single, 90-minute session, the research team presented an overview of the study, and then a consultation manager from the Center for Community Health led discussion including poll questions to quantify attitudes on topics such as preferred study terminology and potential motivators for participation. Open-ended questions encouraged panel members to share their personal experiences, concerns, and suggestions. This feedback provided a broader understanding of perceptions regarding inpatient EEG and delirium research. Key insights from the panel guided the subsequent redesign of our recruitment protocol, as described in the Results, to align with patient and caregiver priorities.

### Analysis

Enrollment rates during defined time periods before and after interventions were calculated as the number of patients enrolled divided by the number of patients approached. We used Chi-square tests with a significance level of 0.05 to compare enrollment rates across different phases of protocol adaptations. Demographic characteristics of enrolled participants before and after adaptations were analyzed using appropriate statistical methods, using Kruskall-Wallis tests for quantitative variables and Chi-square tests for categorical variables, or Fisher exact tests when observed or expected counts were less than five for a cell. Statistical analyses were performed in Python (version 3.12) using the SciPy library (version 1.15.0).

## Results

### Baseline rates of enrollment

Over the first year of the study (September 5, 2023 – August 9, 2024), we approached 367 patients, with 68 (18.5%) consenting to participate. Baseline enrollment rates revealed a difference between services, with neurologic services having a higher enrollment rate (50.0%, 31/62) compared to non-neurologic services (12.1%, 37/305). Furthermore, patients older than 65 years old, who are most vulnerable to delirium, had lower enrollment (12.7%, 37/292) than younger patients (41.3%, 31/75). Given a predetermined target of 20% enrollment for older adults on non-neurologic services, these differing rates raised the question as to what factors might contribute to participation in an observational wearable EEG delirium study, including whether the major concerns were regarding wearable devices or the personal relevance of the study.

### Reasons for not enrolling

Logistical considerations, such as anticipated discharge or inability to reach a LAR, accounted for a portion of non-enrollments (30.1%, Table 1). Additional logistical barriers included interruption by clinical care (9.7%, 29/299), unexpected language barriers (0.7%, 2/299), and cross-enrollment in a competing study (0.3%, 1/299). However, the largest group of non-enrollments, consisting of patients and surrogate decision makers (51.2%), declined participation due to personal or situational reasons, most commonly described as “too much going on” (15.4%, 46/299), requests to be approached by research team at a later time (7.4%, 22/299), lack of interest (5.7%, 17/299), discomfort or fatigue related to illness (5.0%, 15/299), preference to wait for family input (2.7%, 8/299), discomfort with research participation (2.3%, 7/299), reluctance to answer daily questions (2.0%, 6/299), or lack of perceived benefit (2.0%, 6/299). Notably, concerns about the EEG devices reflected only a small proportion of reasons for not enrolling (8.7%, 26/299). These findings suggested a need to understand reasons for participation in more detail to enhance recruitment among broader potential participants.

**Table 1.** Reasons for not participating before interventions.

| Reason for Not Enrolling | n=299 | % |
| --- | --- | --- |
| **Patient/Surrogate Expressed Reason** | **153** | **51.2%** |
| Too much going on | 46 | 15.4% |
| EEG concerns (Hair, discomfort, shower) | 26 | 8.7% |
| Asked to come back later | 22 | 7.4% |
| No interest | 17 | 5.7% |
| Too much discomfort (due to illness)/too tired | 15 | 5.0% |
| Wanted to wait for family | 8 | 2.7% |
| Uncomfortable with research | 7 | 2.3% |
| Don't want to answer questions every day | 6 | 2.0% |
| Did not see benefit in participation (personal or altruistic) | 6 | 2.0% |
| **Logistical Reasons** | **90** | **30.1%** |
| Possible discharge soon | 58 | 19.4% |
| Interrupted by clinical care | 29 | 9.7% |
| Unexpected language barrier | 2 | 0.7% |
| Cross-enrolled in another study (competing study) | 1 | 0.3% |
| **Need for Surrogate** | **56** | **18.7%** |
| Could not reach LAR | 40 | 13.4% |
| Unexpectedly did not have capacity to consent | 16 | 5.4% |

The primary reasons for non-enrollment were collected via open-ended questioning of patients and surrogate decision-makers. Responses were subsequently reviewed and categorized by the research team using thematic analysis. Percents reflect the number of patients/surrogates who identified that factor divided by the total number of patients/surrogates declining participation.

### Community Panel

To understand patient attitudes more deeply and clarify the expressed vague reasons for not enrolling, we convened a community panel with seven individuals who had either been hospitalized or served as a primary caregiver within the past five years. Of the six participants who provided demographic data, all were Black/African American females, with an age distribution spanning early adulthood to late middle age (18–24: n=2; 45–54: n=2; 55–64: n=2). Six panel members responded to poll questions, offering varied opinions on the most helpful terminology to describe the study, showing greater support for framing the study around “brain resilience” or “the impact of being sick on the brain” rather than using the word “delirium” (Figure 1A-B). Most panel members (5/6) felt that educational materials and information about delirium would be more motivating than financial incentives (1/6) or the potential for their cognitive data to be used for clinical care (2/6). Half said that personal health information for their own review, such as sleep data, would motivate them to participate (3/6, Figure 1C). Mirroring the small proportion of hospitalized patients identifying EEG as a specific deterrent to enrollment, only one panel member identified wearing an EEG device as a significant deterrent, in contrast to more panel members identifying other optional research procedures, such as blood draws (4/6) or video recordings (4/6), as deterrents (Figure 1D). Several panel members specifically highlighted personal connections to delirium as reasons to participate in such a study: one had experienced post-operative delirium, another discussed their hospitalization experiences more generally, and two others had family members with dementia and recognized symptoms that were described in the overview.

**Figure 1.**
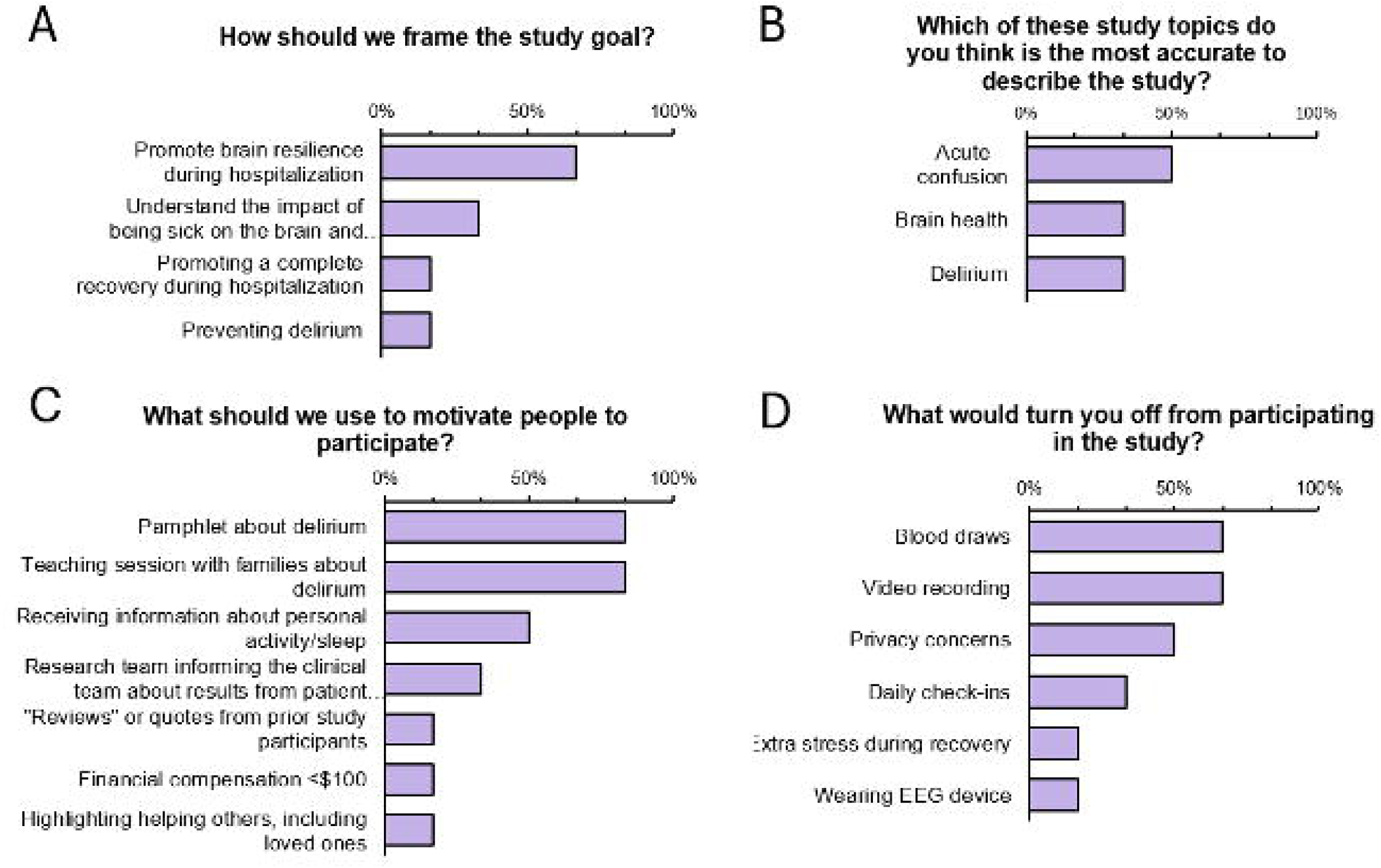
Surveys from community panel. Six community panel participants responded to various poll questions during the panel. Participants indicated the terminology that appealed to them most for describing (A) the study goal and (B) the study topic. (C) Participants were asked to select all the listed recruitment interventions they thought would motivate them or other patients to participate in the SABRE study. The one participant who selected “other” suggested highlighting how their participation could help others in the future, especially their children. (D) Participants selected all the factors that would significantly turn them off from participating in the study.

### First protocol adaptation: Personalization of recruitment conversations

Insights from the community panel guided two major recruitment protocol changes. The first intervention, implemented August 9, 2024, involved tailoring recruitment conversations to patients’ experiences. Recruiters personalized the verbal study introduction to each patient to emphasize delirium symptoms relevant to the individual. This was done by evaluating the medical record and discussing with nurses if the patient had been experiencing any common delirium symptoms (e.g., sleep disturbance, difficulty concentrating, decreased energy). Since we noticed that sleep disturbance was a common experience, whenever no clear delirium symptom was identified for potential participant, we defaulted to highlighting common hospital-related sleep concerns. This personalized symptom based discussion resulted in a significant increase in overall enrollment rates, from 18.5% (68/367) to 30.9% (58/188, χ^2^(1) = 10.8, p = 0.001, Figure 2). The enrollment rate for patients on non-neurologic services increased from 12.1% (37/305) to 29.8% (54/181, χ^2^(1) = 23.4, p<0.001) and those for people over 65 years old increased from 12.7% (37/292) to 30.6% (49/160, χ^2^(1) = 21.6, p < 0.001) (Figure 2B-C).

**Figure 2.**
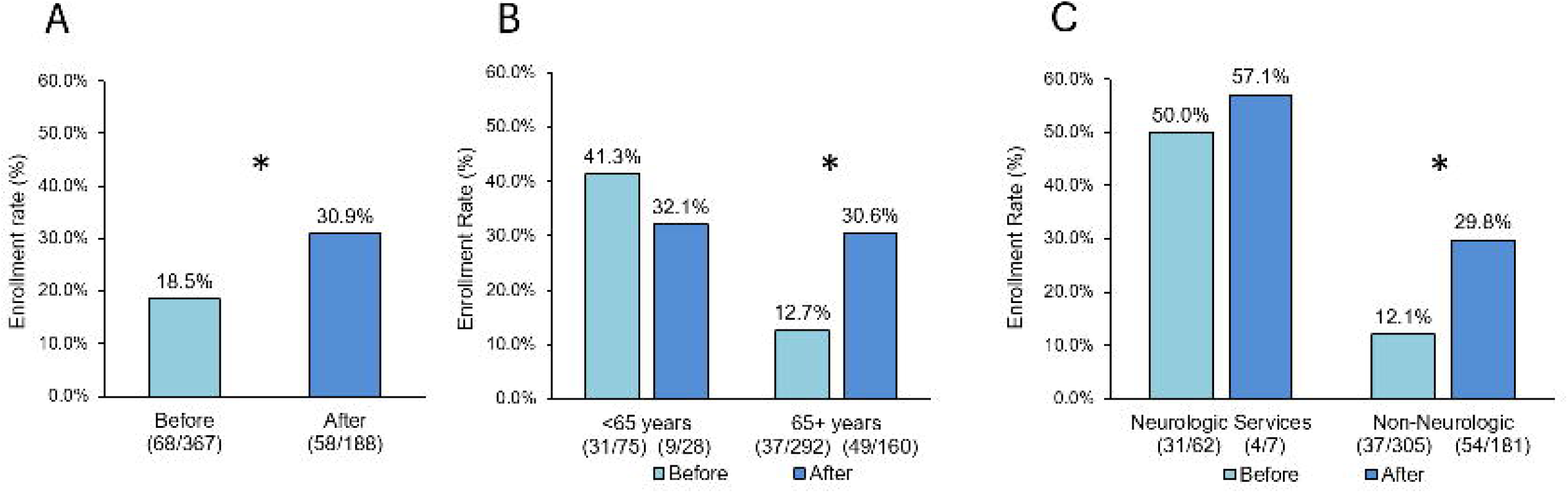
Rates of enrollment before and after protocol changes. (A) Overall enrollment rates, Before and After recruitment protocol changes. (B) Enrollment rates of patients younger than compared to 65 years and older, Before and After protocol changes. (C) Rates of enrollment for patients on neurologic or non-neurologic services, Before and After recruitment protocol changes. Comparisons were made using chi-square tests, * indicates before and after comparison has a p value of ≤0.001.

Following protocol adaptations, we asked a subset of patients why they enrolled in the study. Interest in sleep was the most common reason for enrolling reported by study participants (16/25, 64%, Table 2). These responses reflected the importance of personal connection to symptoms, such as sleep disturbance or confusion, and having a personal interest in the research being done.

**Table 2.** Expressed reasons for enrolling in the research study.

| **Motivating factors** | **n** | **%** |
| --- | --- | --- |
| Interest in sleep | 16 | 64% |
| Interest in research or healthcare | 4 | 16% |
| Interest in confusion | 4 | 16% |
| Experience with delirium | 3 | 12% |
| Nothing else to do | 3 | 12% |
| Interest in effects of infection | 2 | 8% |
| Altruism | 1 | 4% |

Reasons for non-enrollment were collected via open-ended questioning of a subset of patients and surrogate decision-makers of patients (n=25) enrolled after protocol changes. Responses were subsequently reviewed and categorized by the research team using thematic analysis. Multiple responses were permitted; therefore, counts reflect the number of patients/surrogates endorsing each factor, and percentages sum to greater than 100%.

### Second protocol adaptation: Introduction of educational materials

Our second recruitment protocol adaptation, implemented on December 16, 2024, introduced two handouts: a double-sided, tri-fold brochure introducing the study; and a 12-page educational booklet about delirium. The study brochure outlined the study’s purpose, procedures, and reasons to participate. The highlighted benefits of the study included learning about brain health during recovery, contributing to research that could help others, and ease of participation. Altruism was a central message, as the brochure cover read, “We need your help to preserve and improve brain health during and after hospitalization.” The educational booklet aimed to educate patients, family, and caregivers about clinical delirium, and provided educational information, prompts for reflection, and suggestions for conversations between patients, families, friends, and the medical team. It was developed based on expert opinion and consensus guidelines, and reviewed by multiple physicians with delirium, hospitalist, and other psychiatric expertise. Compared to the symptom personalization protocol change (47/148 enrolled), offering educational materials did not further increase enrollment rates (11/40, χ²(1) = 0.27, p = 0.61) for this study, though they were anecdotally appreciated by those who enrolled.

### Demographics

Table 3 summarizes the demographic characteristics of participants enrolled before and after the recruitment adaptations. After protocol adaptations, the median age of enrolled participants increased from 69.0 to 75.5 years (Kruskal-Wallis test, p < 0.001) and patients were more likely to be enrolled from general medical services (Fisher exact test, p < 0.001) Gender, race, ethnicity and rates of baseline cognitive impairment, were largely similar across patients who enrolled before and after adaptations. Additionally, comorbidities and admission diagnosis classifications were statistically similar across patients who enrolled before and after adaptations, with a wide distribution of admission diagnoses (Supplementary Table S1).

**Table 3.**
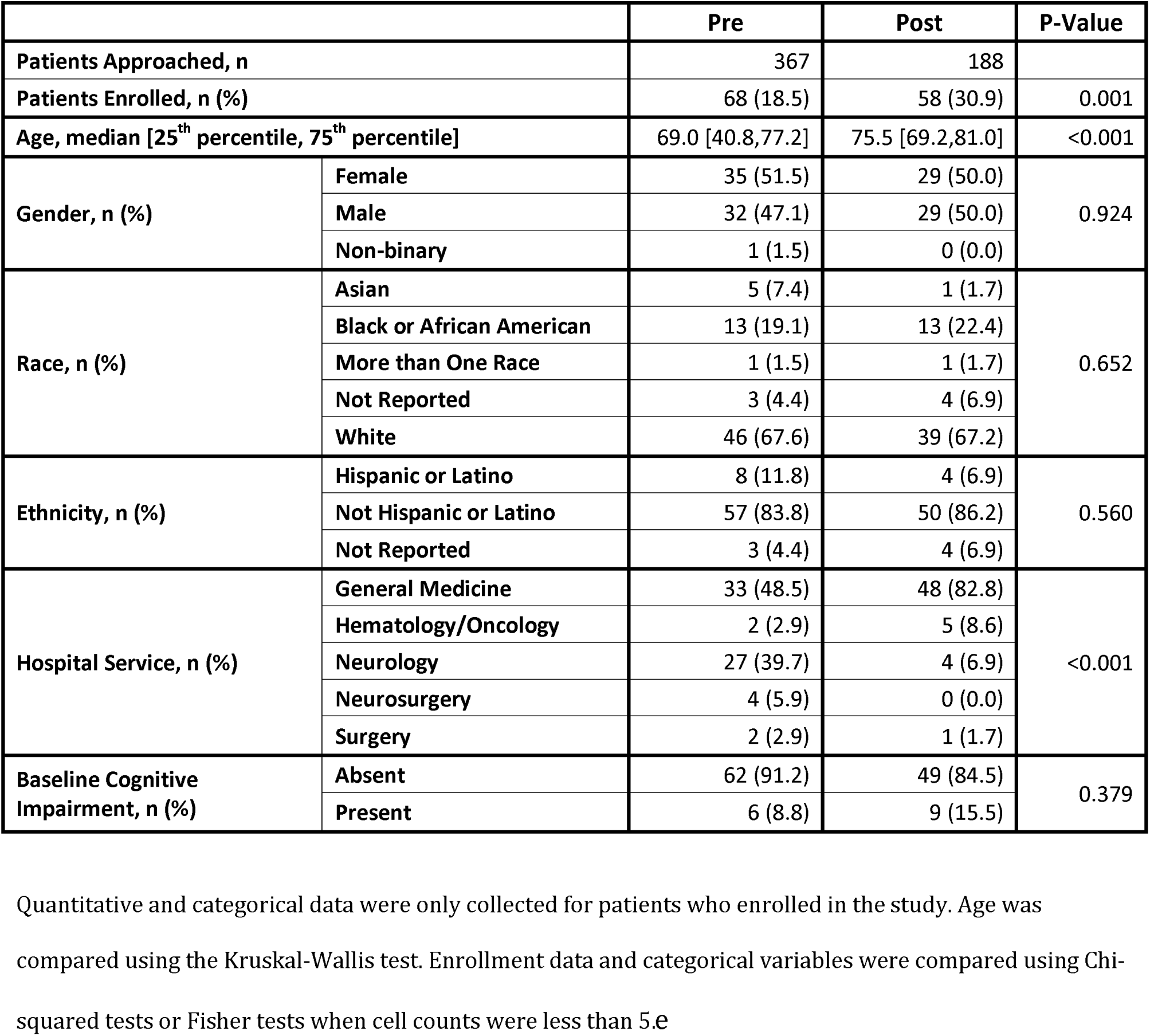
Characteristics of patients who enrolled in the study before and after interventions.

|  |  | Pre | Post | P-Value |
| --- | --- | --- | --- | --- |
| **Patients Approached, n** |  | 367 | 188 |  |
| **Patients Enrolled, n (%)** |  | 68 (18.5) | 58 (30.9) | 0.001 |
| **Age, median [25<sup>th</sup> percentile, 75<sup>th</sup> percentile]** |  | 69.0 [40.8,77.2] | 75.5 [69.2,81.0] | <0.001 |
| **Gender, n (%)** | **Female** | 35 (51.5) | 29 (50.0) | 0.924 |
|  | **Male** | 32 (47.1) | 29 (50.0) |  |
|  | **Non-binary** | 1 (1.5) | 0 (0.0) |  |
| **Race, n (%)** | **Asian** | 5 (7.4) | 1 (1.7) | 0.652 |
|  | **Black or African American** | 13 (19.1) | 13 (22.4) |  |
|  | **More than One Race** | 1 (1.5) | 1 (1.7) |  |
|  | **Not Reported** | 3 (4.4) | 4 (6.9) |  |
|  | **White** | 46 (67.6) | 39 (67.2) |  |
| **Ethnicity, n (%)** | **Hispanic or Latino** | 8 (11.8) | 4 (6.9) | 0.560 |
|  | **Not Hispanic or Latino** | 57 (83.8) | 50 (86.2) |  |
|  | **Not Reported** | 3 (4.4) | 4 (6.9) |  |
| **Hospital Service, n (%)** | **General Medicine** | 33 (48.5) | 48 (82.8) | <0.001 |
|  | **Hematology/Oncology** | 2 (2.9) | 5 (8.6) |  |
|  | **Neurology** | 27 (39.7) | 4 (6.9) |  |
|  | **Neurosurgery** | 4 (5.9) | 0 (0.0) |  |
|  | **Surgery** | 2 (2.9) | 1 (1.7) |  |
| **Baseline Cognitive Impairment, n (%)** | **Absent** | 62 (91.2) | 49 (84.5) | 0.379 |
|  | **Present** | 6 (8.8) | 9 (15.5) |  |

Quantitative and categorical data were only collected for patients who enrolled in the study. Age was compared using the Kruskal-Wallis test. Enrollment data and categorical variables were compared using Chi-squared tests or Fisher tests when cell counts were less than 5.e

Although we could not compare patients who enrolled to those who did not enroll, as patients who did not enroll did not provide consent to collect personal data, we compared patients who enrolled in the study to all patients admitted to the hospital during this time who met inclusion criteria. Patients who enrolled were in the study were older, had higher rates of baseline cognitive impairment than the broader population, and were more likely to be on medical and neurology services than the general admission population (Supplementary Table S2). Patients who enrolled in the study were also more likely to have a history of cerebrovascular disease, chronic pulmonary disease, dementia, peripheral vascular disease, and rheumatic disease than the broader population, and less likely to have a history of malignancy (Supplementary Table S3).

## Conclusions

### Overall Findings and Interpretation

Symptom-based personalization of recruitment significantly increased enrollment in an inpatient wearable EEG delirium study, particularly in older patients and those on non-neurological services. In the context of prospective delirium research, it was not the presence of potentially uncomfortable elements that initially kept enrollment rates low but rather the lack of perceived relevance or personal interest. The addition of educational materials about delirium and study participation did not further affect patients’ likelihood of enrolling.

### Wearable devices were not a major deterrent

Contrary to our original concerns, we found that the wearable EEG device was not a clear deterrent to participation. Existing literature shows that older adults are more inclined to use a wearable health device if it addresses a present health need.^20^ For instance, experiencing multiple falls increased the perceived benefit of a fall tracker.^20^ We concluded that the perceived benefits and interest in the study topic were initially insufficient to outweigh any perceived burden of an inpatient study, but that this was potentially modifiable.^21^ Across inpatient research involving older adults, recruitment success may be improved not only by mitigating study procedure burden but moreso by highlighting the personal relevance of the research topic to the patient’s immediate health experience.

### Input from the Community

To clarify the sometimes nebulous, vague, or non-specific reasons for not enrolling, we hosted a community panel to explore perspectives and attitudes toward inpatient delirium research. Panel members illuminated the importance of ensuring that the recruitment process sparks patients’ interest and connects with their priorities. For panel members, this interest overwhelmed hypothetical deterrents from the study and were more important than financial compensation. Half of panel members recommended providing personal health information as an incentive to participate, for example information about sleep in the hospital. However, validated sleep metrics unfortunately do not yet exist for hospitalized older adults, especially those with delirium. Community input on patient-centered outcomes can help researchers prioritize development efforts, however. Findings from the panel ultimately implied that an interest in research, interest in the study’s focus on brain resilience, and a personal stake in the study goal would incentivize participation. We were able to modify the recruitment protocol by incorporating these patient and caregiver perspectives.

### Importance of immediate relevance

Previous literature supports the idea that when patients do not perceive a study as helping them clinically, which is the case for observational research, it is more effective to appeal to their intrinsic interest in the study topic.^22^ However, patients without delirium but at risk of developing it may underestimate future risks^23^ and cannot be expected to have the same investment in an unfamiliar condition they may develop compared to more active clinical issues they currently experience. Hoping to improve delirium knowledge among approached patients, we reviewed the previous literature^3^ and spoke with community panel members for recommendations, which led us to provide written information on the study and on delirium. Our findings show that providing informational materials based on expert and consensus guidelines did not further increase enrollment. It is possible that the materials would benefit from further community involvement, but it appears more likely that while educational materials may improve general understanding of delirium, they may not alter patients’ perceptions or priorities within the short timeframe in which inpatient enrollment decisions are made. While our informational materials about delirium and the SABRE study did not significantly increase enrollment, there is likely value in investigating this approach for other research topics. In acute inpatient research settings, potential participants may have limited time and face stressors and distractions that can hinder their ability to engage with educational materials. We hypothesize that such materials may be more effective in less stressful settings where the enrollment process can unfold over a longer duration.

### How to think about altruism in the context of research participation

An additional component of the written materials was to highlight altruistic benefits of participating in research, but this did not significantly increase enrollment further. This finding was unexpected based on previous literature that reports altruism as the primary motivator for participants of clinical trials and observational research.^22,24^ Altruistic motivation has been linked to a perceived semi-personal benefit of helping one’s own community, such as those with the same medical condition,^25^ and may be bolstered by an empathic understanding of the importance of what is being studied. Recruiting a broad group of participants on varied medical services may have resulted in a more heterogeneous population than rare or identity-based conditions and may not foster a strong sense of group affiliation or in-group empathy.^26^ Without that identity, altruism may be a weaker motivator.^27^

### Study Population

Patients with neurologic conditions were initially more likely to enroll than those with non-neurologic diagnoses, possibly reflecting their greater familiarity with EEG monitoring and its relevance to their care. Adapting the recruitment protocol to emphasize symptoms of delirium relevant to individual patients broadened enrollment of the study’s population to older adults, who frequently have lower rates of enrollment in clinical research studies.^3^ The protocol adaptation increased the enrollment rate for patients with non-neurologic conditions to a greater degree than for those with neurologic conditions, demonstrating that symptom-based recruitment conversations can effectively broaden participation by helping studies connect with patients in personally meaningful ways.

### Sleep as a motivating factor

For patients who did not experience cognitive symptoms, sleep served as an effective default topic to attract patients to the study. Most people are personally familiar with sleep disturbance, especially during hospitalization.^28^ Hospitalized patients frequently report poor sleep or suffer from insomnia and circadian rhythm disorders. Primary causes for poor sleep while hospitalized include discomfort due to illness and the unfamiliar, chaotic hospital environment. Poor sleep can worsen patients’ subjective wellness, perception of quality of care, emotional health, and physical outcomes during hospitalization, so a study addressing sleep disturbance was more pertinent to hospitalized patients’ immediate experiences^28^ than one couched in the broader clinical terminology of delirium, in which sleep and circadian dysfunction is nearly universally disrupted.^29^ Although sleep was a particularly relevant symptom for many participants in our inpatient delirium study, we suggest that identifying patient centered symptoms personally relevant to prospective participants may be a generalizable strategy to increase engagement in other prospective research studies.

### Limitations

This study has several limitations. First, reasons for not enrolling were self-reported and often vague, limiting the depth of understanding of specific deterrents among the broader inpatient population. For this reason, we performed a community panel to gain deeper insight into concerns. The qualitative insights from the community panel may be subject to self-selection bias, as participants likely possessed a pre-existing interest in brain health or clinical research. Furthermore, the small sample size and the absence of formal data regarding education, health literacy, and socioeconomic status among panel members may limit the contextual depth and broad generalizability of these findings. However, we believe these insights remained highly relevant to our target population because all panel members were recruited from the immediate community surrounding the academic medical center where the study was conducted, ensuring a shared geographic and institutional connection. Regarding the study’s quantitative results, our sample size varied for different phases of the recruitment protocol. We approached fewer patients after adaptations were implemented (n=188) compared to the number approached before (n=367), in part due to increased success enrolling patients over the timeframe of the study. Moreover, while we tracked enrollment rate changes resulting from personalized approach conversations, we added the informational and educational resources 130 days after this change, which limited our ability to understand the independent value of informational and educational resources, and may have been affected by a ceiling effect from improvements already achieved. The order of addition was limited by the time to obtain IRB approval for the new documents. Lastly, while we observed a strong association, given the before-and-after design of this study, other unmeasured factors might have influenced participation over time.

### Future directions

The need to establish a personal connection and interest in potential research for potential participants is an actionable insight to improve participation in inpatient research in older patients. By focusing on symptoms personally relevant to patients, researchers can significantly improve enrollment in studied of multifaceted clinical syndromes, such as delirium, to engage broader patient populations.

## Data Availability

All data produced in the present study are available upon reasonable request to the authors

## Acknowledgements

The corresponding author affirms that they listed everyone who contributed significantly to the work and obtained written consent from all contributors who are not authors and are named in the Acknowledgment section. The authors wish to acknowledge the community panels members, Josefina Serrato of the Northwestern University Center for Community Health for guidance in the Shared Resource Panel with community panel members, and Drake Gorecki for draft review.

## Author Contributions

Study Concept and Design (EYK), Acquisition of Participants and Data (NC, SD, CZ, IS), Analysis and interpretation of data (NC, EYK), and Preparation of manuscript (NC, EYK)

