## Supplement for "A Personalized, Symptom-Based Approach to Boost Research Participation in Hospitalized Older Adults"

*Supplementary Figure S 1* EEG wearable device

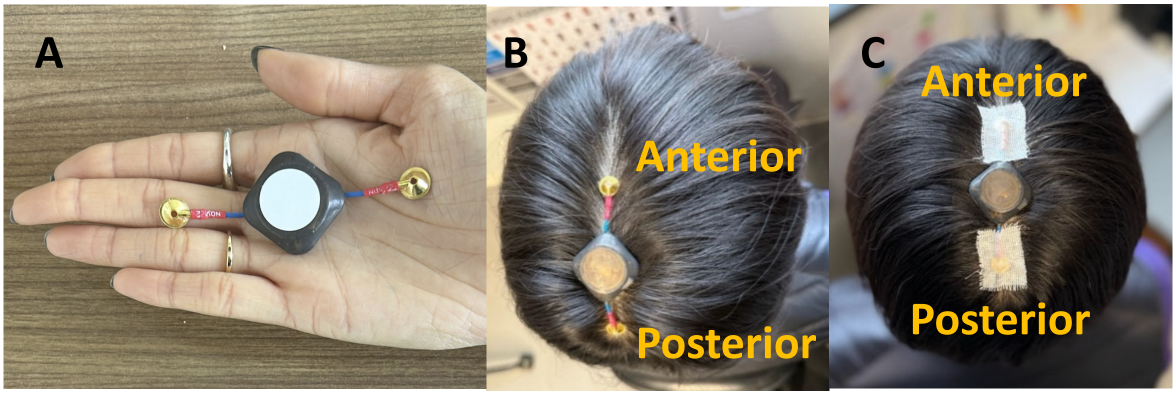

**A** The EEG device is approximately 25mm along each edge with gold cups attached at diagonal corners for placement at the vertex of the head.

**B** The device is shown at the vertex of the head with the anterior electrode at standard International EEG electrode location Cz, and with the posterior electrode 7cm behind at Pz.

**C** The device is affixed to the head using standard EEG application procedures including collodion and cotton gauze.

*Supplementary Table S 1* Comorbidities and admission diagnoses of patients who enrolled in the study before and after interventions.

| | Pre (n=68) | Post (n=58) | P-Value |
|---|---|---|---|
| **Charlson Comorbidities, n (%)** | | | |
| AIDS HIV | 0 (0.0) | 0 (0.0) | 1.000 |
| Cerebrovascular disease | 17 (25.0) | 18 (31.0) | 0.579 |
| Chronic pulmonary disease | 16 (23.5) | 21 (36.2) | 0.173 |
| Congestive heart failure | 14 (20.6) | 18 (31.0) | 0.255 |
| Dementia | 4 (5.9) | 4 (6.9) | 1.000 |
| Diabetes with chronic complications | 7 (10.3) | 6 (10.3) | 1.000 |
| Diabetes without chronic complications | 11 (16.2) | 12 (20.7) | 0.673 |
| Hemiplegia or paraplegia | 1 (1.5) | 1 (1.7) | 1.000 |
| Liver disease | 5 (7.4) | 7 (12.1) | 0.552 |
| Malignancy | 11 (16.2) | 14 (24.1) | 0.372 |
| Metastatic Solid Tumor | 4 (5.9) | 3 (5.2) | 1.000 |
| Myocardial infarction | 2 (2.9) | 0 (0.0) | 0.497 |
| Peptic ulcer disease | 3 (4.4) | 2 (3.4) | 1.000 |
| Peripheral vascular disease | 17 (25.0) | 20 (34.5) | 0.333 |
| Renal disease | 13 (19.1) | 12 (20.7) | 1.000 |
| Rheumatic disease | 6 (8.8) | 7 (12.1) | 0.762 |
| **Admission WHO ICD-10 Diagnosis Chapters, n (%)** | | | |
| Certain infectious and parasitic diseases | 1 (1.5) | 3 (5.2) | 0.294 |
| Codes for special purposes | 0 (0.0) | 1 (1.7) | |
| Diseases of the blood and blood-forming organs and certain disorders involving the immune mechanism | 2 (2.9) | 1 (1.7) | |
| Diseases of the circulatory system | 11 (16.2) | 9 (15.5) | |
| Diseases of the digestive system | 1 (1.5) | 1 (1.7) | |
| Diseases of the eye and adnexa | 1 (1.5) | 0 (0.0) | |
| Diseases of the genitourinary system | 3 (4.4) | 5 (8.6) | |
| Diseases of the musculoskeletal system and connective tissue | 5 (7.4) | 1 (1.7) | |
| Diseases of the nervous system | 15 (22.1) | 4 (6.9) | |
| Diseases of the respiratory system | 3 (4.4) | 6 (10.3) | |
| Endocrine, nutritional and metabolic diseases | 2 (2.9) | 5 (8.6) | |
| Factors influencing health status and contact with health services | 2 (2.9) | 2 (3.4) | |
| Mental and behavioural disorders | 2 (2.9) | 3 (5.2) | |
| Neoplasms | 4 (5.9) | 2 (3.4) | |
| Symptoms, signs and abnormal clinical and laboratory findings, not elsewhere classified | 16 (23.5) | 15 (25.9) | |

Charlson comorbidities were extracted from the electronic medical record and compared using Chi-squared tests or Fisher tests when expected or actual cell counts were less than 5. P-values are not corrected for multiple comparisons given the exploratory and descriptive nature of these data. Admission diagnoses were extracted from the electronic medical record and classified according to World Health Organization (WHO) International Classification of Disease-10 (ICD-10) diagnosis chapters. A Fisher test was used to compare the distribution of diagnoses between the two groups.

*Supplementary Table S 2* Comparison of demographics of enrolled patients to all potentially eligible patients admitted during the study period.

| | | Enrolled | All Admits | P-Value |
|---|---|---|---|---|
| **Patients n** | | 126 | 7702 | |
| **Age, median [25th percentile, 75th percentile]** | | 72.0 [62.0,80.0] | 62.0 [47.0,72.0] | <0.001 |
| **Gender, n (%)** | **Female** | 64 (50.8) | 3694 (48.0) | 0.357 |
| | **Male** | 61 (48.4) | 3978 (51.6) | |
| | **Non-binary** | 1 (0.8) | 30 (0.4) | |
| **Race, n (%)** | **American Indian or Alaska Native** | 0 (0.0) | 17 (0.2) | 0.056 |
| | **Asian** | 6 (4.8) | 271 (3.5) | |
| | **Black or African American** | 26 (20.6) | 1419 (18.4) | |
| | **More than One Race** | 2 (1.6) | 39 (0.5) | |
| | **Native Hawaiian or Other Pacific Islander** | 0 (0.0) | 11 (0.1) | |
| | **Not Reported** | 7 (5.6) | 1054 (13.7) | |
| | **White** | 85 (67.5) | 4891 (63.5) | |
| **Ethnicity, n (%)** | **Hispanic or Latino** | 12 (9.5) | 569 (7.4) | 0.464 |
| | **Not Hispanic or Latino** | 107 (84.9) | 6536 (84.9) | |
| | **Not Reported** | 7 (5.6) | 597 (7.8) | |
| **Hospital Service, n (%)** | **General Medicine** | 81 (64.3) | 2795 (36.3) | <0.001 |
| | **Hematology/Oncology** | 7 (5.6) | 1439 (18.7) | |
| | **Neurology** | 31 (24.6) | 876 (11.4) | |
| | **Neurosurgery** | 4 (3.2) | 348 (4.5) | |
| | **Surgery** | 3 (2.4) | 2244 (29.1) | |
| **Baseline Cognitive Impairment, n (%)** | **Absent** | 111 (88.1) | 7451 (96.7) | <0.001 |
| | **Present** | 15 (11.9) | 251 (3.3) | |

Quantitative and categorical data were compared between patients who enrolled vs. all eligible patients admitted during the study period. Age was compared using the Kruskal-Wallis test. Enrollment data and categorical variables were compared using Chi-squared tests or Fisher tests when cell counts were less than 5.

*Supplementary Table S 3* Comorbidities and admission diagnoses of enrolled patients to all potentially eligible patients admitted during the study period.

| | Enrolled (n=126) | All Admits (n=7702) | P-Value |
|---|---|---|---|
| **Charlson Comorbidities, n (%)** | | | |
| AIDS HIV | 0 (0.0) | 57 (0.7) | 1.000 |
| Cerebrovascular disease | 35 (27.8) | 1161 (15.1) | <0.001 |
| Chronic pulmonary disease | 37 (29.4) | 1528 (19.8) | 0.011 |
| Congestive heart failure | 32 (25.4) | 1733 (22.5) | 0.507 |
| Dementia | 8 (6.3) | 95 (1.2) | <0.001 |
| Diabetes with chronic complications | 13 (10.3) | 711 (9.2) | 0.793 |
| Diabetes without chronic complications | 23 (18.3) | 1529 (19.9) | 0.739 |
| Hemiplegia or paraplegia | 2 (1.6) | 181 (2.4) | 1.000 |
| Liver disease | 12 (9.5) | 1167 (15.2) | 0.104 |
| Malignancy | 25 (19.8) | 2464 (32.0) | 0.005 |
| Metastatic Solid Tumor | 7 (5.6) | 878 (11.4) | 0.056 |
| Myocardial infarction | 2 (1.6) | 328 (4.3) | 0.209 |
| Peptic ulcer disease | 5 (4.0) | 206 (2.7) | 0.389 |
| Peripheral vascular disease | 37 (29.4) | 1549 (20.1) | 0.014 |
| Renal disease | 25 (19.8) | 1762 (22.9) | 0.485 |
| Rheumatic disease | 13 (10.3) | 388 (5.0) | 0.014 |
| **Admission WHO ICD-10 Diagnosis Chapters, n (%)** | | | |
| Certain infectious and parasitic diseases | 4 (3.2) | 60 (0.8) | <0.001 |
| Codes for special purposes | 1 (0.8) | 6 (0.1) | |
| Congenital malformations, deformations and chromosomal abnormalities | 0 (0.0) | 24 (0.3) | |
| Diseases of the blood and blood-forming organs and certain disorders involving the immune mechanism | 3 (2.4) | 143 (1.9) | |
| Diseases of the circulatory system | 20 (15.9) | 1287 (16.7) | |
| Diseases of the digestive system | 2 (1.6) | 568 (7.4) | |
| Diseases of the ear and mastoid process | 0 (0.0) | 10 (0.1) | |
| Diseases of the eye and adnexa | 1 (0.8) | 9 (0.1) | |
| Diseases of the genitourinary system | 8 (6.3) | 250 (3.2) | |
| Diseases of the musculoskeletal system and connective tissue | 6 (4.8) | 203 (2.6) | |
| Diseases of the nervous system | 19 (15.1) | 472 (6.1) | |
| Diseases of the respiratory system | 9 (7.1) | 206 (2.7) | |
| Diseases of the skin and subcutaneous tissue | 0 (0.0) | 65 (0.8) | |
| Endocrine, nutritional and metabolic diseases | 7 (5.6) | 190 (2.5) | |
| External causes of morbidity and mortality | 0 (0.0) | 16 (0.2) | |
| Factors influencing health status and contact with health services | 4 (3.2) | 1458 (18.9) | |
| Injury, poisoning and certain other consequences of external causes | 0 (0.0) | 341 (4.4) | |

| | | |
|---|---|---|
| Mental and behavioural disorders | 5 (4.0) | 78 (1.0) |
| Neoplasms | 6 (4.8) | 945 (12.3) |
| Other | 0 (0.0) | 506 (6.6) |

Charlson comorbidities were extracted from the electronic medical record and compared using Chi-squared tests or Fisher tests when expected or actual cell counts were less than 5. P-values are not corrected for multiple comparisons given the exploratory and descriptive nature of these data. Admission diagnoses were extracted from the electronic medical record and classified according to World Health Organization (WHO) International Classification of Disease-10 (ICD-10) diagnosis chapters. A Fisher test was used to compare the distribution of diagnoses between the two groups.
